# A case of autosomal dominant osteopetrosis type II with a severe bone phenotype but no amino acid converting mutation in the *CLCN7* gene

**DOI:** 10.1101/2021.07.08.21257202

**Authors:** Jochen G. Hofstaetter, Gerald J. Atkins, Masakazu Kogawa, Stéphane Blouin, Barbara M. Misof, Paul Roschger, Andreas Evdokiou, L. Bogdan Solomon, David M. Findlay, Nobuaki Ito

## Abstract

Autosomal Dominant Osteopetrosis type II (ADOII), also known as Albers-Schönberg disease, is caused by mutation of the *CLCN7* chloride channel gene and is characterized by reduced bone resorption. Here we report on an individual with the classic features of ADOII, who had a history of fractures from childhood, displayed high bone mass and characteristic “sandwich vertebrae” on x-ray. Our genetic analyses showed no amino acid converting mutation in the patient’s DNA but we did find evidence of haploinsufficiency of *CLCN7* mRNA. An iliac crest bone sample from the patient revealed bone tissue and material abnormalities relative to normal controls based on quantitative backscattered electron imaging and histomorphometric analyses. Additionally to lamellar bone, we observed significant amounts of woven bone and mineralised cartilage, as well as an increased frequency and thickness (up to 15 microns) of cement lines. Giant osteoclasts with numerous nuclei were present. The bone mineralisation density distribution (BMDD) of the entire bone area revealed markedly increased average mineral content of the dense bone (CaMean T-score +10.1) and frequency of bone with highest mineral content (CaHigh T-score +19.6), suggesting continued mineral accumulation and lack of bone remodelling. Osteocyte lacunae sections (OLS) characteristics were unremarkable except the OLS shape which was unusually circular. Together, our findings suggest that the reduced expression of *CLCN7* mRNA in osteoclasts, and possibly also osteocytes, causes poorly remodelled bone with abnormal bone matrix with high mineral content. This together with the lack of adequate bone repair mechanisms makes the material brittle and prone to fracture.

## INTRODUCTION

Osteopetrosis is a group of rare heterogeneous hereditary diseases characterized by increased bone mass, which is caused by reduced bone resorption, due to reduced numbers and or impaired function of osteoclasts [1]. Autosomal dominant osteopetrosis type II (ADOII) (MIM# 166600), also known as Albers-Schönberg disease, is the most frequent (1:20.000) form of osteopetrosis [2] and is mostly caused by heterozygous dominant negative mutations of the gene encoding chloride channel, voltage-sensitive 7 (*CLCN7*; OMIM #602727) [3, 4]. The autosomal recessive form (ARO) (MIM# 611490) is less frequent [4, 5]. The protein product of *CLCN7* has an important role in mature osteoclasts, extruding chloride from the ruffled boarder to facilitate proton extrusion by V-ATPase [5-7], thus providing the ability to acidify the extracellular environment. Interestingly, osteopetrotic conditions based on *CLCN7* mutations show great variation in their presentation and severity [3].

ADOII individuals attain average height and have normal life expectancy but x-rays reveal high bone mass and characteristic “sandwich vertebrae” [1]. Despite the dramatic increase in Dual-energy X-ray Absorptiometry (DXA)-derived areal bone mineral density (BMD), ADOII individuals have a high risk of sustaining low energy fractures. We report here a case of a middle-aged male of normal height, who at surgery for a subtrochanteric fragility fracture of the femur was found to have unusually high bone mass. Consequently, genetic analysis of the *CLCN7* gene was performed and a transiliac bone biopsy sample was taken for examinations of histological and material abnormalities using light microscopy and scanning electron microscopy. In particular, we have successfully used quantitative backscattered electron imaging (qBEI) for the characterization of the bone mineralization phenotype in patients with genetic diseases, thus providing insights into the genotype-bone phenotype relationship [8-10].

## MATERIALS & METHODS

### Clinical description of patient

Research ethics approval for the study was obtained from the Royal Adelaide Hospital Human Research Ethics Committee (Approval No. RAH130114) and informed consent to both conduct the study and publish the findings was obtained from the proband. The proband was the second child of non-consanguineous parents. Several of the proband’s relatives had at least some features of ADOII including radiographic hallmarks of typical ‘sandwich’ vertebrae but had an inconsistent history of low trauma fracture (Fig. 1A). Further access to the clinical history is available on request from the corresponding author.

**Figure 1:**
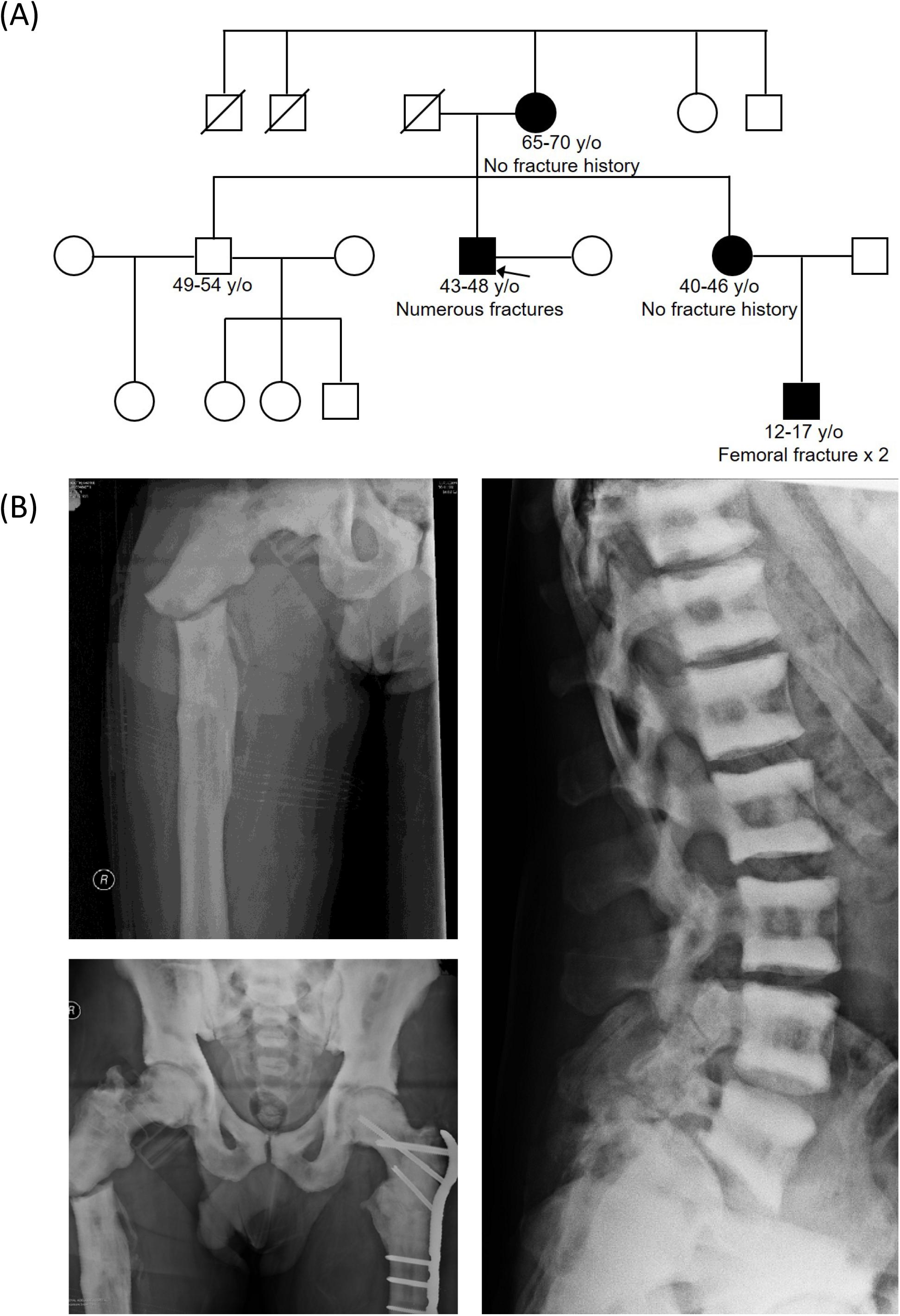
(A) The proband’s pedigree, showing male and female relatives who apparently share the same genetic abnormality. (B) Radiographs of the proband, showing the right proximal femoral fracture, the previously treated left femoral fracture, the extremely radio-dense skeleton and the ‘sandwich’ vertebrae, typical of ADOII.

The proband reported no problems with dentition, is an occasional smoker with a history of moderate alcohol consumption. He was clinically diagnosed with ADOII/Albers-Schönberg disease in childhood following a fracture based on his distinctive x-ray images. He suffered numerous fractures throughout his childhood and adulthood at multiple skeletal sites, mainly from relatively low impact traumas. At ages 42-47 in separate incidents, he suffered fractures to his left proximal femur, which fractured again after several months and then his right proximal femur (Fig. 1B). Plain radiographs revealed a remarkably high bone mass throughout his body, with ‘sandwich’ vertebrae (Fig. 1B). Blood tests showed no evidence of anemia, leukocytopaenia or other abnormalities. He underwent surgery where it was found that the femur could not be nailed because of insufficient endosteal space and that the hardness of the cortical bone made drilling screw holes and plating difficult. A bone biopsy sample was obtained from the right iliac crest at the time of surgery for examination of histological and bone material quality aspects. Genetic analysis and an osteoclastogenesis assay were also performed.

### Sequencing

Genomic DNA (gDNA) and total RNA were isolated from peripheral blood mononuclear cells (PBMC). Total RNA and complementary DNA (cDNA) were prepared, as described previously [11]. gDNA and complementary DNA (cDNA) sequencing for *CLCN7*, including splice sites and 5′, 3′ untranslated regions (5′, 3′ UTR) in the proband DNA were sequenced using Applied Biosystems 3730 and 3730xl sequencers (Applied Biosystems, Carlsbad, CA, USA). Quantitative PCR was performed for *CLCN7* and glyceraldehyde-3-phosphate dehydrogenase *(GAPDH)* mRNA, using RNA extracted from PBMC of the proband and a healthy control, as described previously [11].

### Osteoclastogenesis assay

Peripheral blood mononuclear cells (PBMC; 5.4 × 10^5^ cells/cm^2^) were incubated in αMEM medium containing added recombinant human (rh)M-CSF (25 ng/ml) and rhRANKL (100 ng/ml). Media were replaced every 3 days thereafter. Osteoclast number was assessed by TRAP staining after 6–9 days, as described previously [12].

### Iliac crest bone biopsy

#### Sample preparation

Commonly used Bordier transiliac biopsy could not be performed due to the increased hardness of the patient’s bone material. Therefore, a diamond drill was used to drill two small holes into the iliac crest and subsequently, a bone chip was obtained from the iliac crest. This bone sample was immediately fixed in 70% ethanol, dehydrated in a graded series of ethanol and embedded undecalcified in polymethylmethacrylate. Subsequently, 3 µm sections for histology/histomorphometry were prepared with a hard tissue microtome (Leica SM2500, Nussloch, Germany). The surface of the residual sample block was prepared by grinding and polishing (Logitech PM5, Glasgow, Scotland) and carbon coated (Agar SEM Carbon Coater; Agar Scientific, Stansted, UK) for quantitative backscattered electron imaging (qBEI) evaluation.

#### Histological assessment

As the bone biopsy sample showed predominantly mineralised tissue and contained almost no free bone marrow space, we could not quantify standard histomorphometric parameters. However, analysis of qBEI overview images of the sample allowed us to determine mineralised tissue volume per tissue volume (corresponding to bone volume per tissue volume, BV/TV). The Goldner’s trichrome or Giemsa stained bone sections were imaged in a light microscope (Axiophot, equipped with digital camera AxioCam HRc, Zeiss, Oberkochen, Germany). The histologic sections were used to assess the presence and appearance of osteoclasts and to discriminate areas of lamellar bone and woven bone using polarized light microscopy. The presence of residual mineralised cartilaginous matrix could be quantified by qBEI based on its non-fibrillar micro-texture and higher mineral content. Cement line phenotyping as well as osteocyte lacunae characteristics were also performed on basis of qBEI (following).

#### Quantitative backscattered electron imaging (qBEI)

qBEI analysis of bone tissue is based on the fact, that the grey levels in the images (if adequately calibrated) are proportional to the local weight percentage of calcium (Ca) in the bone matrix as described elsewhere [13]. We acquired qBEI images in Zeiss DSM 962 and Zeiss Supra 40 instruments (both Zeiss, Oberkochen, Germany) for quantification of global and local calcium content within the biopsy sample, including the bone mineralisation density distribution (BMDD), the BV/TV, tissue details like mineralised cartilage, cement lines and osteocyte lacunae sections (OLS). Both instruments were equipped with a four-quadrant semiconductor backscatter electron detector and operated at 20 kV accelerating voltage for the beam electrons and a scan speed of 100 seconds per frame.

In order to determine the patient’s BMDD the Zeiss DSM 962 was operated at 110 pA probe current and 15 mm working distance. A series of images with a pixel resolution of 3.6 μm from about 19 mm^2^ of the iliac crest bone area were recorded. Grey level histograms were derived from the qBEI images representing frequency distributions of pixels with a certain Ca content (Ca weight%), denominated BMDD. Five parameters were deduced from this BMDD: The weighted mean Ca-concentration of the bone area (CaMean), the peak position of the histogram (CaPeak, indicating the most frequent Ca concentration), the full width at half maximum of the distribution (CaWidth, indicative for the heterogeneity in matrix mineralisation), the percentage of bone areas having a Ca-concentration lower than 17.68 wt% Ca (CaLow), and the percentage of bone areas having a Ca-concentration higher than 25.30 wt% Ca (CaHigh, corresponding to fully mineralised bone areas, mainly interstitial bone and cement lines). These parameters were compared to reference values for cancellous BMDD for adult individuals [14].

For the assessment of BV/TV, a binary image (discriminating between bone marrow and mineralised matrix) was generated from the entire sectioned bone area. Discrimination between bone marrow space and bone area was performed by a at fixed grey level threshold corresponding to 0.87 weight % Ca. Image analysis and quantification of the two areas were performed by custom made routines using the software ImageJ [15].

In order to study mineralised tissue details, the Zeiss Supra 40 operating at about 280 pA probe current, 10 mm working distance and a pixel resolution in the range of 1.76 up to 0.25 µm, was used. Additionally, energy dispersive X-ray (EDX) measurements for semi-quantitative information on elemental composition were performed on small areas of mineralised bone, cartilage, and cement lines using an EDX system coupled to the SEM SUPRA 40 and equipped with an EDS Silicon Driftdetector (X-Max, Oxford Instrument, UK). The energy of the beam electrons was adjusted to 10 KeV and the EDX spectra were analysed based on Oxford INCA software.

The OLS analysis is a 2D-characterization of osteocyte lacunae sections based on qBEI images with a pixel resolution of 0.88 μm, which were subsequently transformed to binary images (distinguishing mineralised from unmineralised bone) using a threshold based on a fixed calcium value of 5.2 wt% Ca [16] (mineralised cartilage areas were excluded for OLS-analysis). The OLS were extracted in these images using a size range between 5 µm^2^ and 200 µm^2^, respectively, and analyzed for the OLS-density (the number of OLS per mineralised bone area), the OLS-porosity (the total area of OLS given as percentage of the total bone area), the mean OLS-area, the mean OLS-perimeter, and the mean OLS-aspect ratio (the major axis over the minor axis of the fitted ellipse) based on a custom-made macro in ImageJ software. OLS-aspect ratio=1 indicates a circle and increasing values indicate increasingly elongated shape of the OLS. Details of assessment of OLS-characteristics are described elsewhere [17]. The presence and viability of the osteocytes within the lacunae cannot be evaluated by this method. The patient’s OLS-characteristics were compared with those measured in transiliac bone biopsy samples from two healthy adult women published previously [18].

## RESULTS

### No amino acid converting mutation in the *CLCN7* gene

Because mutation of the *CLCN7* chloride channel gene is known to be causative for ADOII, we performed whole gene sequencing of *CLCN7*, using genomic DNA (gDNA) extracted from the proband’s peripheral blood monocytes. We focused on the coding sequence, together with splice sites and predicted 5′ and 3′ UTRs of *CLCN7* gene. No amino acid converting mutations could be found in the proband’s *CLCN7* gene sequence. However, two different heterozygous single nucleotide polymorphisms (SNP) were detected in exon 1 (c.126T > C/T, rs3751884) and exon 14 (c. 1170A > A/T) (Fig. 2A). The presence of these two SNPs (each existing in almost 50% of the population) allowed us to investigate haploinsufficiency of *CLCN7* mRNA expression. Sequencing of exon 1 and exon 14 cDNA revealed that *CLCN7* mRNA is only transcribed from one allele in the proband (Fig. 2A). Furthermore, quantitative PCR for *CLCN7* corrected by *GAPDH* levels showed that the expression level of *CLCN7* mRNA is less than one third of that of a healthy control (Fig. 2B).

**Figure 2:**
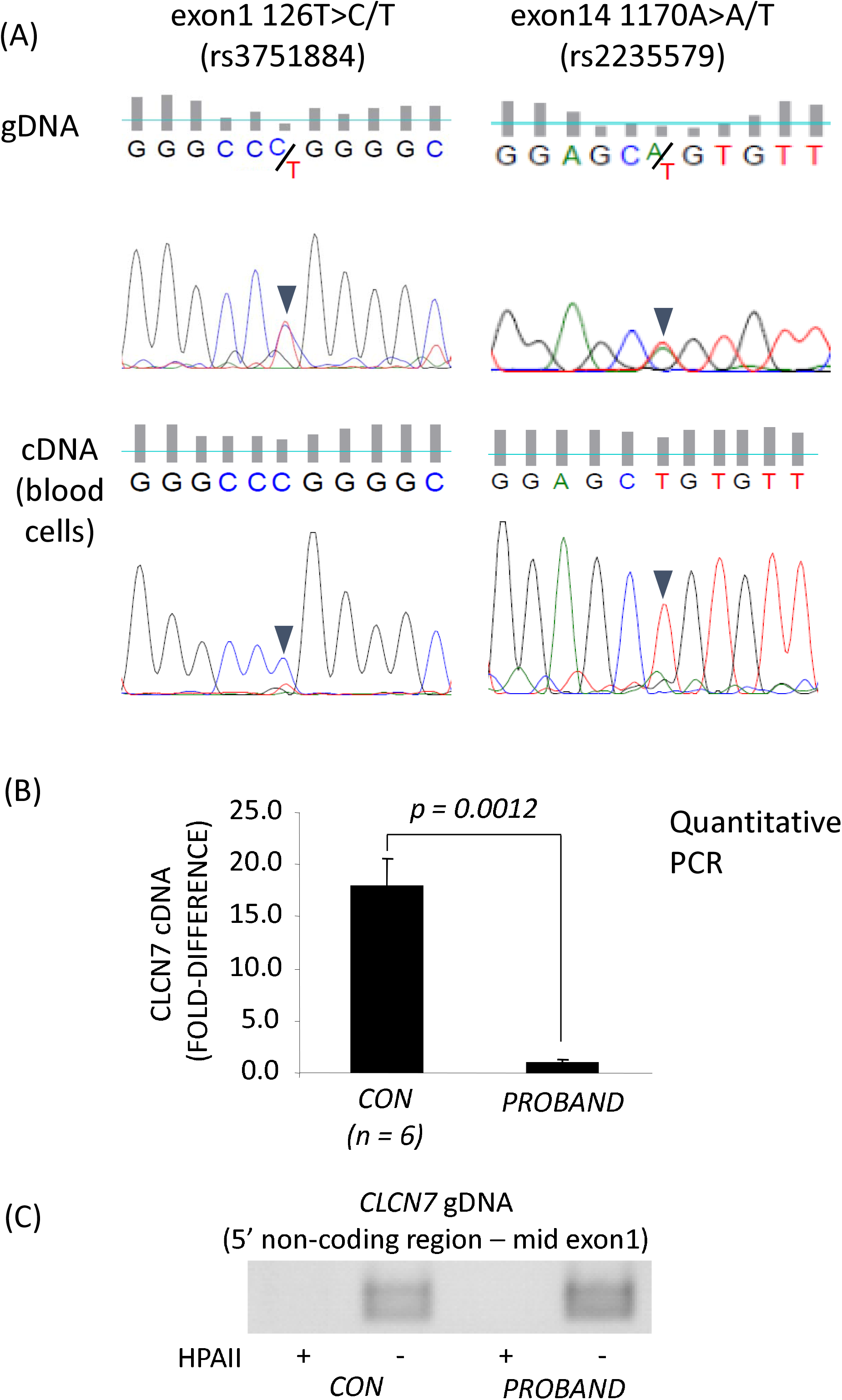
(A) Evidence for two different heterozygous single nucleotide polymorphisms (SNP) were found in exon 1 (c.126T > C/T, rs3751884) and exon 14 (c. 1170A > A/T). (B) Quantitative PCR for *CLCN7* normalised to *GAPDH* expression showed that the expression level of *CLCN7* mRNA is less than one third of that of a healthy control.

### Intact ability to differentiate from monocytes to osteoclasts

To investigate the possibility that the proband had a deficiency in osteoclastogenesis, we performed an *in vitro* analysis. The proband’s PBMC retained the ability to differentiate into large multinucleated TRAP-positive osteoclast-like cells when cultured with exogenous rhRANKL and rhM-CSF (Supplementary Fig. S1), suggesting that osteoclast formation was unimpaired.

### Abnormalities in phenotype of bone histology and mineralization

#### Histology

Images of histological staining are shown in Figure 3. Most remarkable, the transiliac bone sample from our patient did not contain typical trabecular features within the marrow space. The tissue volume was almost completely filled with bone material (Fig. 4A) with BV/TV of 95.5 % and 84.6 % in regions R1 and R2, respectively.

**Figure 3:**
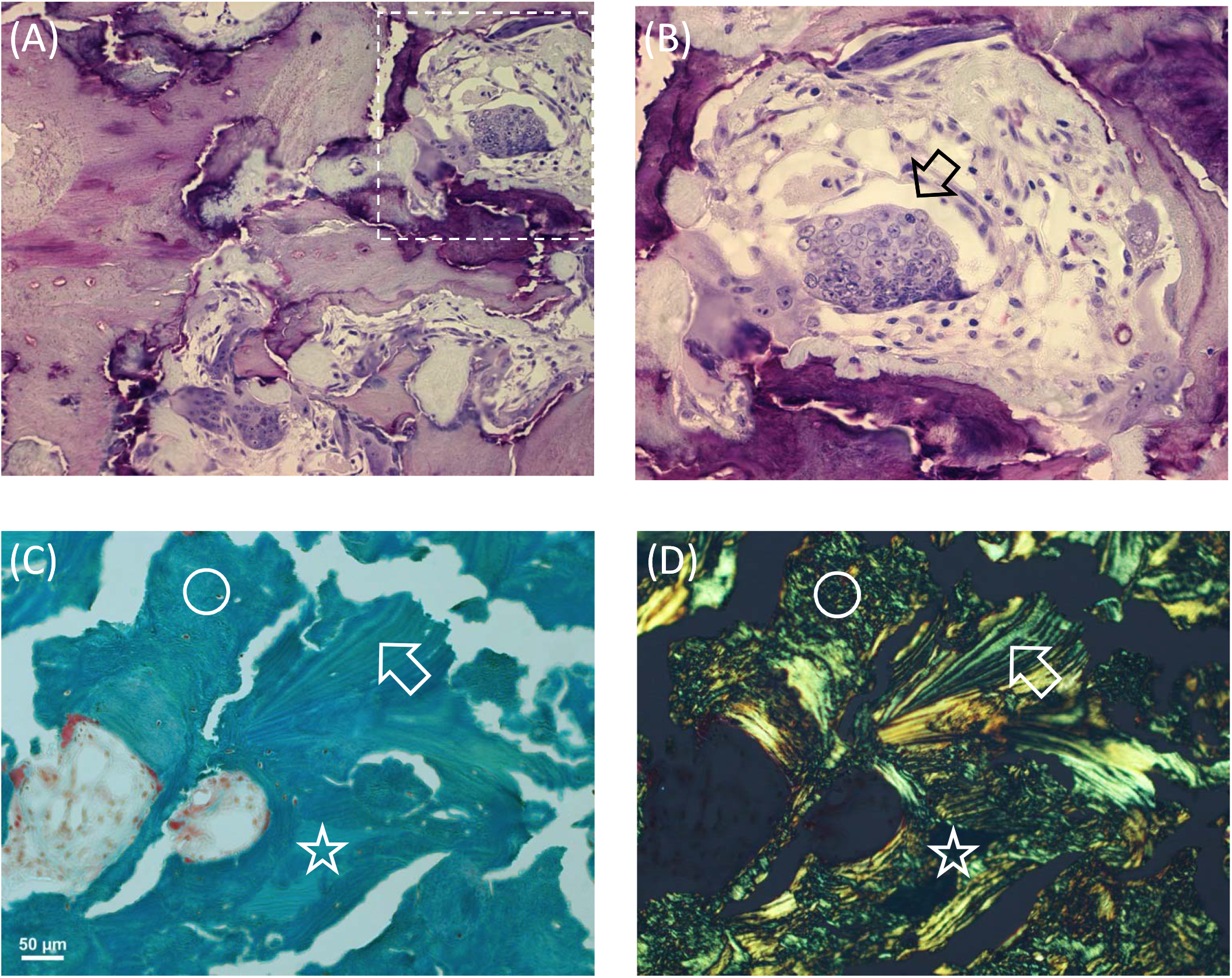
Histologic evaluation of the bone biopsy. (A), (B) Giemsa-stained bone sections under light microscopy. Image in (B) is a detail of (A) (indicated by the dashed area) revealing a giant osteoclast (arrow) with a high number of nuclei. (C), (D) Goldner-stained section under normal light microscopy (C) and under polarized light (D). The images give evidence for the presence of lamellar bone (arrow), woven bone (circle) and mineralised cartilage (asterisk) as frequently found in the patient’s iliac crest sample.

**Figure 4:**
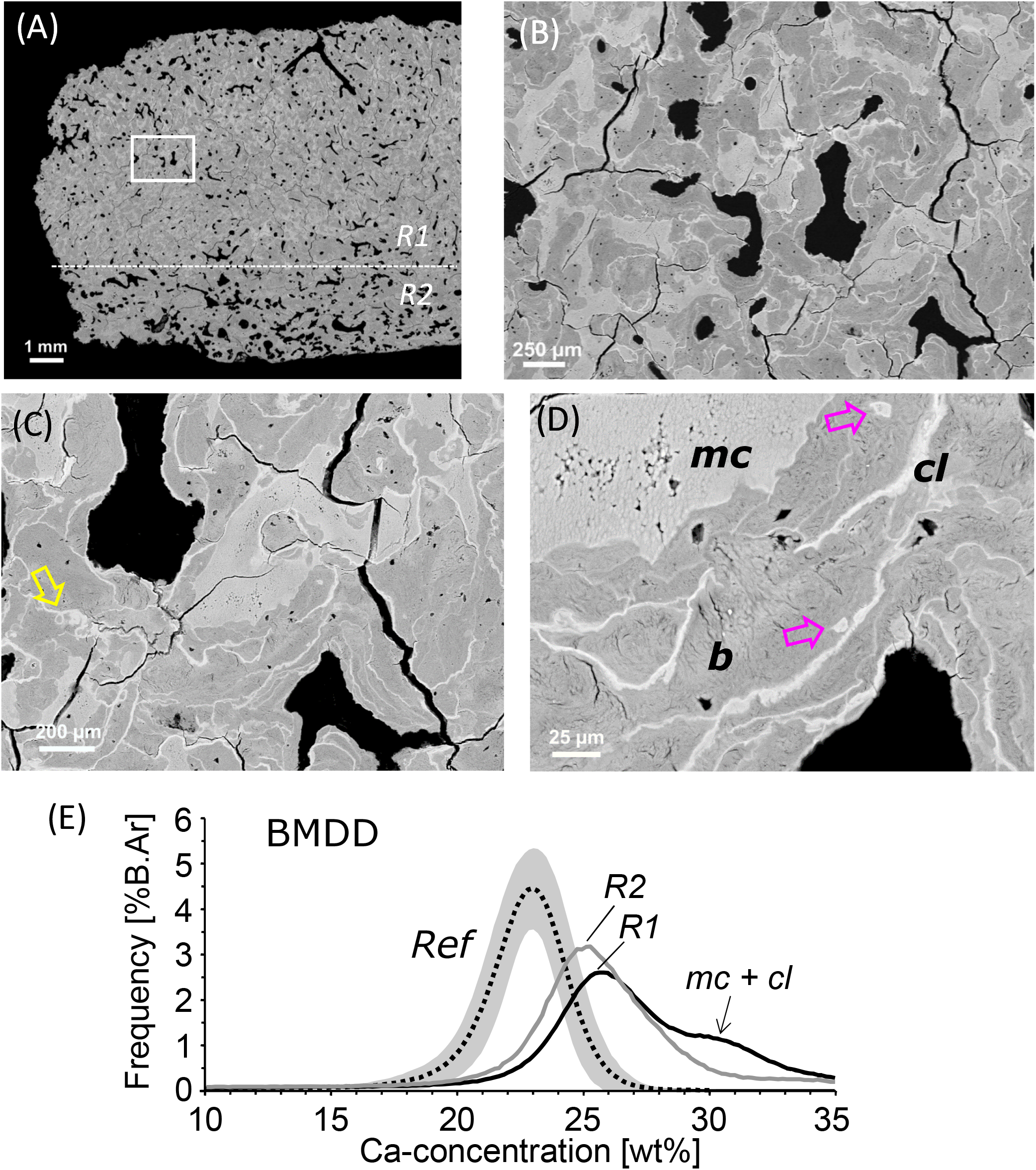
qBEI overview image of the proband’s iliac bone biopsy sample (A), showing zones of differential mineral density (shades of grey-white). A dense region R1 nearly completely filled with bone matrix, and a less dense region R2 can be distinguished. The corresponding bone mineralisation density distribution (BMDD) histograms within these zones are shown in (E). (B)-(D) show the area indicated by the rectangular box at higher magnifications. Clearly visible are the areas of mineralised cartilage (mc) and the cement lines (cl) by their brighter grey levels compared to bone (b), areas indicated in (D). Of note is the unusual structure of cement lines with small loops (yellow arrow in C) and kinks, as well as mineralised osteocyte lacunae (pink arrows in D). (E) The BMDD of the proband’s bone at both regions R1 and R2 is clearly shifted to higher calcium concentrations compared to trabecular bone from healthy adults (Reference BMDD, mean and 95^th^ CI indicated by dotted line and grey area, respectively). The “shoulder” (marked by the arrow) reflects the contribution of mineralised cartilage and cement lines to the BMDD.

Despite of the compactness of bone tissue, typical secondary osteons (with concentric bone lamellae) could only rarely be observed. The bone sample contained canals consistent with primary osteons. Giemsa staining gave evidence for the presence of a mixture of bone and cartilage (Fig. 3A). By setting a threshold of 27.9 weight % Ca in the qBEI images (Fig. 4) for demarcation of mineralized cartilage (mineral content beyond threshold) from bone (below threshold), the amount of mineralized cartilage was found to be 32.3 % of the mineralized tissue area in region 1 (R1 in Fig. 4A). Consistent with our observation of apparent exuberant osteoclast differentiation from the proband’s PBMC, giemsa-stained sections showed the presence of giant osteoclasts with numerous nuclei (Fig. 3B). Goldner-stained sections examined by polarized light microscopy showed that the bone sample was only partly lamellar (collagenous fibrils arranged into regular lamellae), whereas another part consisted of woven bone (disorganized fibrillar structures) (Fig. 3C, 3D).

Noticeable also was the extremely high number of cement lines per bone area as visualized by qBEI (Fig. 4). We could distinguish between two different types of cement lines: thin with normal or increased mineral content and thick with extremely high mineral content (Fig. 4, Fig. 5). These highly mineralised cement lines had a thickness up to 15 micrometer and were unusual in their appearance as they contained frequently small loops and kinks (Fig. 4B-D, Fig. 5). A line profile of Ca content through a region of cartilage, bone and cement line clearly demonstrated the extraordinary high mineral content of these cement lines (up to 37 weight % Ca corresponding to 93 weight % hydroxyapatite mineral) (Fig. 5). Moreover, elemental EDX analysis revealed that exclusively in these cement lines detectable amounts of the element Fluor (0.5 to 1.1 atom %) were present. Furthermore, EDX analysis of the Ca to P ratios showed that the mineral present in mineralised cartilage, bone and cement lines all had a similar calcium-to-phosphorus ratio of 1.71 to 1.76, which is close to the theoretical value of 1.66 of pure hydroxyapatite.

**Figure 5:**
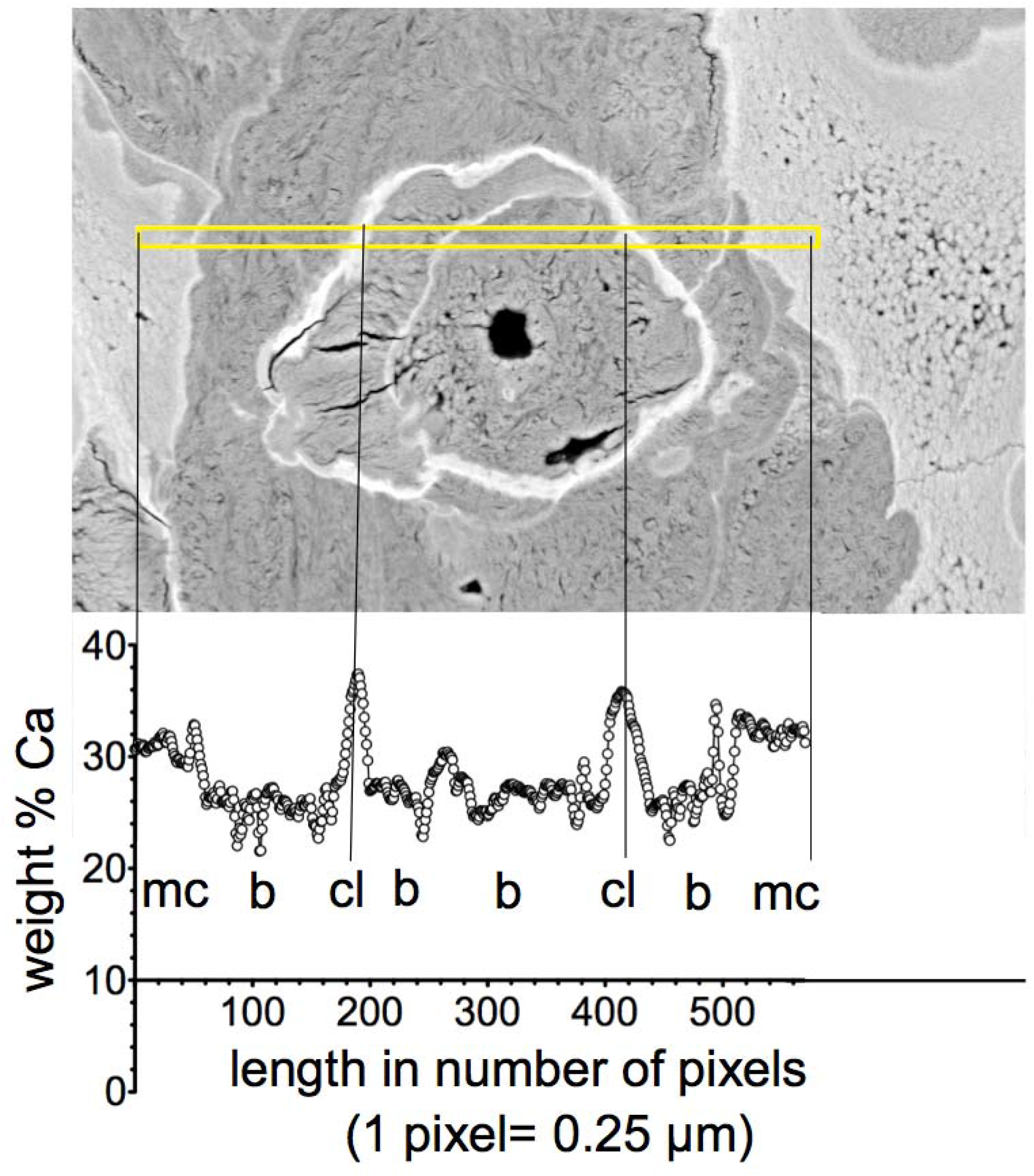
Line profile (yellow indicated line area) of calcium concentration through a tissue area containing mineralised cartilage (mc), bone (b) and thick cement lines (cl).

#### OLS

For the OLS-characteristics, 1446 osteocyte lacunae sections from the mineralised bone area (mineralised cartilage was excluded for this analysis) were analyzed, yielding an OLS-porosity of 0.34%, OLS-density of 159 per mm^2^, OLS-area of 21.27μm^2^, and OLS-perimeter of 19.06μm, which were all within the range observed in two control samples. However, the proband’s OLS-aspect ratio of 2.09 was lower compared to those from the controls (2.62 and 2.54) indicating more roundish osteocyte lacunae.

#### BMDD

The BMDD from our patient was separately measured in the dense region R1 and in the less dense region R2 (see Fig. 4A, 4E). At both sites, the BMDD was distinctly shifted to higher calcium concentrations and broadened compared to the reference BMDD from healthy adult individuals (Fig. 4E). Consequently, the BMDD parameter values were clearly different from reference data (for BMDD outcomes and T-scores from both regions R1 and R2; see Table 1). The average (CaMean T-scores +10.1 and +6.7 for regions R1 and R2, respectively) and most frequent calcium concentrations (CaPeak T-scores +7.4 and +5.6), the heterogeneity of mineralisation (CaWidth T-scores +4.9 and +4.4), as well as the percentage of bone with highest mineral content (CaHigh T-scores +19.6 and +14.6), were highly elevated, while the percentage of low mineralised areas (CaLow T-scores -1.1 and -0.4) was reduced.

**Table 1:** Comparison of the BMDD-parameters between the osteopetrotic bone biopsy sample and reference data base of healthy adults [14].

| BMDD | Reference<br>(n=52) | Patient with osteopetrosis |  |  |  |
| --- | --- | --- | --- | --- | --- |
|  |  | Region R1 | R1 T-score | Region R2 | R2 T-score |
| Ca<sub>MEAN</sub><br>[wt% Ca] | 22.20 ± 0.45 | 26.74 | +10.1 | 25.21 | +6.7 |
| Ca<sub>PEAK</sub><br>[wt% Ca] | 22.94 ± 0.39 | 25.82 | +7.4 | 25.13 | +5.6 |
| Ca<sub>WIDTH</sub><br>[Δwt% Ca] | 3.35 ± 0.34 | 5.03 | +4.9 | 4.51 | +4.4 |
| Ca<sub>LOW</sub><br>[%B.Ar] | 4.93 ± 1.57 | 3.18 | -1.1 | 4.28 | -0.4 |
| Ca<sub>HIGH</sub><br>[%B.Ar%] | 5.55 ± 3.32 | 70.65 | +19.6 | 54.11 | +14.6 |

R1 = dense bone region, R2 = less dense bone region (as indicated in Fig 5A and 5E). Reference data indicate mean ± SD, the patient's BMDD outcomes are shown additionally by T-scores (based on the given reference data).

## DISCUSSION

Here we describe a patient with profound osteopetrosis, i.e. with extremely elevated bone mass and with vertebrae showing a characteristic “sandwich” structure. The patient had also a history of numerous fractures indicating a high degree in brittleness of the bone material. Approximately 70% of patients with Albert-Schönberg disease (ADOII) have heterozygous dominant negative mutations of the *CLCN7* gene (#166600) [5]. In our patient, no amino acid converting mutation was evident in the *CLCN7* gene but because of the presence of SNPs in exons 1 and 14, we detected haploinsuffiency in the *CLCN7* gene resulting in reduced *CLCN7* mRNA expression, which is likely to be responsible for the observed phenotype. However, the cause of lack in transcription of one *CLCN7* allele in this patient could not be identified. Thus, it is not clear whether haploinsufficiency of *CLCN7* alone can explain ADOII. For example, there is an apparent absence of a bone phenotype in heterozygous family members of ARO patients with a loss of function mutation of *CLCN7*, or in heterozygous *Clcn7*^*+/-*^ mice [6, 7]. It should also be noted that silent, or synonymous, mutations that do not alter the amino acid sequence can nevertheless affect protein function [19]. Hence, it is possible that additional factors contribute to the phenotype in the affected individual described here.

In general, defects in the *CLCN7* gene result in reduced bone resorption by osteoclasts and perhaps disturb the coupling between osteoclastic resorption and bone formation by osteoblasts [20]. At an organ level, this dysfunction of osteoclasts leads to the diffuse sclerotic bone phenotype within the entire skeleton, including the characteristic “sandwich vertebrae” as observed in our patient. In line with the latter, we detected extremely high bone mass at the tissue level and obviously dysfunctional, abnormally shaped giant osteoclasts not necessarily attached to inner bone surfaces, whose large size might be a response to compensate for their defective activity. The mineralized tissue volume was found to be up to 96%, compared to 22% in healthy individuals of the same age [21]. A consequence of the impaired bone (re)modelling in our patient is the persistence of a large fraction of cartilage originating from iliac crest development [22], together with the presence of primary woven bone within mature secondary lamellar bone matrix. Such islands of mineralised cartilage within the bone matrix are a characteristic histologic feature of osteopetrosis [23].

Furthermore, we observed an abnormally high number and increased thickness of cement lines. Generally, cement lines are considered to be the first material deposited onto the resorption surface by mononuclear cells [24]. This resorption surface usually shows subtle undulations reflecting the resorption (Howship) lacunae generated by osteoclasts [25]. In our patient, however, numerous cement lines contained frequent small loops and kinks of unknown origin. We hypothesize that these might originate from the inefficient attempts of the osteoclast to resorb the underlying bone material. Moreover, the thickness of the cement lines in our patient was partly up to 3-fold of the normally observed thickness of 5 microns [25]. In previous histologic assessments of osteopetrotic bone, the presence of an amorphous layer between osteoclasts and the lamellar bone surface was described [23, 26, 27]. Semba et al. [27] reported that this material might contribute to the thickened cement lines in osteopetrosis. In any case, it is well-known that, in general, cement lines are more highly mineralised than the surrounding bone matrix [25, 28], which can be attributed to mineralisation of non-collagenous protein in the former. Noteworthy, in our patient most of the cement lines are mineralized to a much higher degree (up to 93 weight % mineral) than usually as can be seen clearly by their brighter appearance in our qBEI images. However, our elemental analysis showed calcium to phosphorus ratios consistent with the presence of hydroxyapatite-like mineral in these unusually thick cement lines. Interestingly, we also found an accumulation of fluoride in these cement lines but not in mineralised cartilage or bone. The underlying mechanism and significance of this observation remain unclear.

In addition to affecting osteoclast activity, *CLCN7* deficiency could also affect mineral embedded osteocytes, the predominant cell type in bone [29-31]. We have reported the expression of *CLCN7* in mouse and human osteocytes and have shown that it is upregulated in an *in vitro* model of perilacunar remodelling [11], a process that reversibly removes mineral from bone, likely for the dual purposes of calcium homeostasis and to maintain an appropriately mineralised bone matrix [29, 32]. The perilacunar and peri-canalicular bone is considered to be a significant calcium reservoir [33, 34], which might be depleted by osteocytic osteolysis in response to high calcium demand [29, 32, 35]. In turn, such a release of calcium would increase the size of the osteocyte lacunae in qBEI images. *CLCN7* haploinsufficiency in contrast might hamper the ability of the osteocyte to acidify its surrounding matrix, which is needed during osteocytic osteolysis [11]. Thus, in the proband the perilacunar and peri-canalicular reservoir is likely highly loaded with calcium, which might contribute to the overall high calcium concentration found in the bone material. In line with this assumption, while we did not observe abnormalities in terms of the number/density or size of osteocyte lacunae, the proband’s bone contained more circular osteocyte lacunae in 2D sections than expected, which may either be attributed to the presence of woven bone [36], and/or an altered ability of the osteocyte to model its lacunar shape due to a disturbed perilacunar remodelling apparatus. It is noteworthy that several mineralised osteocyte lacunae were also seen in our patient’s bone material reflecting dead osteocytes, which indicate defective bone turnover, repair and impaired bone quality [37].

At the material level, the iliac crest bone biopsy from our patient in general contained greatly increased calcium concentrations and higher heterogeneity of mineralisation compared to reference data from healthy adult individuals. This is in line with the high calcium concentrations in CLCN7-dependent osteopetrosis reported by others [38]. At least two factors are contributing to these increased calcium concentrations in our patient, namely the significant amount of mineralised cartilage, which achieves a higher degree of calcium concentration than bone, and increased bone tissue age due to the disturbed bone turnover, thus leading to prolonged secondary mineralisation. At this point, it should be noted that BMDD reflects the history of bone turnover. New bone matrix laid down by osteoblasts onto the cement-layer of a resorption lacunae starts to mineralize after a matrix maturation of a few days. The further mineral accumulation occurs in a primary rapid phase (up to about 70% of full mineralisation within a few days) and a slower secondary phase (up to full mineralisation within months to years) [39, 40]. Thus, bone is composed of bone packets of different ages with correspondingly different mineral content generating a certain pattern of mineralisation as described by the BMDD. In healthy adult individuals, the BMDD in cancellous bone falls within a relatively narrow range, likely reflecting the degree and distribution of mineral to achieve optimal bone strength and toughness [41]. In contrast, the high mineral content in our patient made the bone material harder, but also more brittle and prone to fracture [42]. In particular, the deviations from uniform lamellar collagen fibril arrangement in the bone matrix by inclusion of woven bone, cartilage and cement lines contribute to a high heterogeneity of the material introduce weak interfaces, which might act as stress concentrators thus promoting initiation of microcracks [43].

In conclusion, we describe an ADOII patient without amino acid converting mutation but with a *CLCN7* haploinsufficiency leading to a remarkably increased bone mass. His iliac crest bone consists of lamellar bone but also of inclusion of mineralised cartilage and primary woven bone with abnormally thick and highly mineralised cement lines. These histologic abnormalities are potentially the result of reduced expression of *CLCN7* in osteoclasts and possibly osteocytes, causing low rates of bone resorption and the lack of internal bone repair, which is the likely cause of his brittle bone, with reduced fracture resistance.

## Data Availability

There are no external datasets related to this paper.

## Disclosures

None of the authors have any disclosures of conflicts of interest to declare.

## Acknowledgements

The authors thank P. Keplinger, S. Lueger, and P. Messmer for technical assistance with sample preparation, light microscopy and qBEI measurements at the Bone Material Laboratory of the Ludwig Boltzmann Institute of Osteology, Vienna, Austria. This work was supported in part by grants from The National Health and Medical Research Council of Australia (NHMRC: ID1047796), the AUVA (Austrian Social Insurance for Occupational Risk) and the OEGK (Austrian Social Health Insurance Fund).

## Authors’ roles

Study design: NI, JGH, GJA and DMF. Study conduct: NI, PR and JGH. Data collection: NI, MK, SB, BMM, PR and JGH. Data analysis: NI, SB, GJA, PR and JGH. Data interpretation: NI, SB, BMM, PR, LBS, DMF, GJA. Drafting manuscript: NI, GJA, DMF. Revising manuscript content: all authors. Approving final version of manuscript: all authors. NI and JGH take responsibility for the integrity of the data analysis.

## FIGURE LEGENDS

**Supplementary Figure S1:** Circulating monocytes from the proband (left) and from a healthy donor (right) are shown. The proband’s cells can differentiate into TRAP positive multinucleated osteoclast-like cells. Cells incubated for 4 days in the presence or absence of rhRANKL are shown, stained for TRAP.

**Supplementary Fig. S1:**
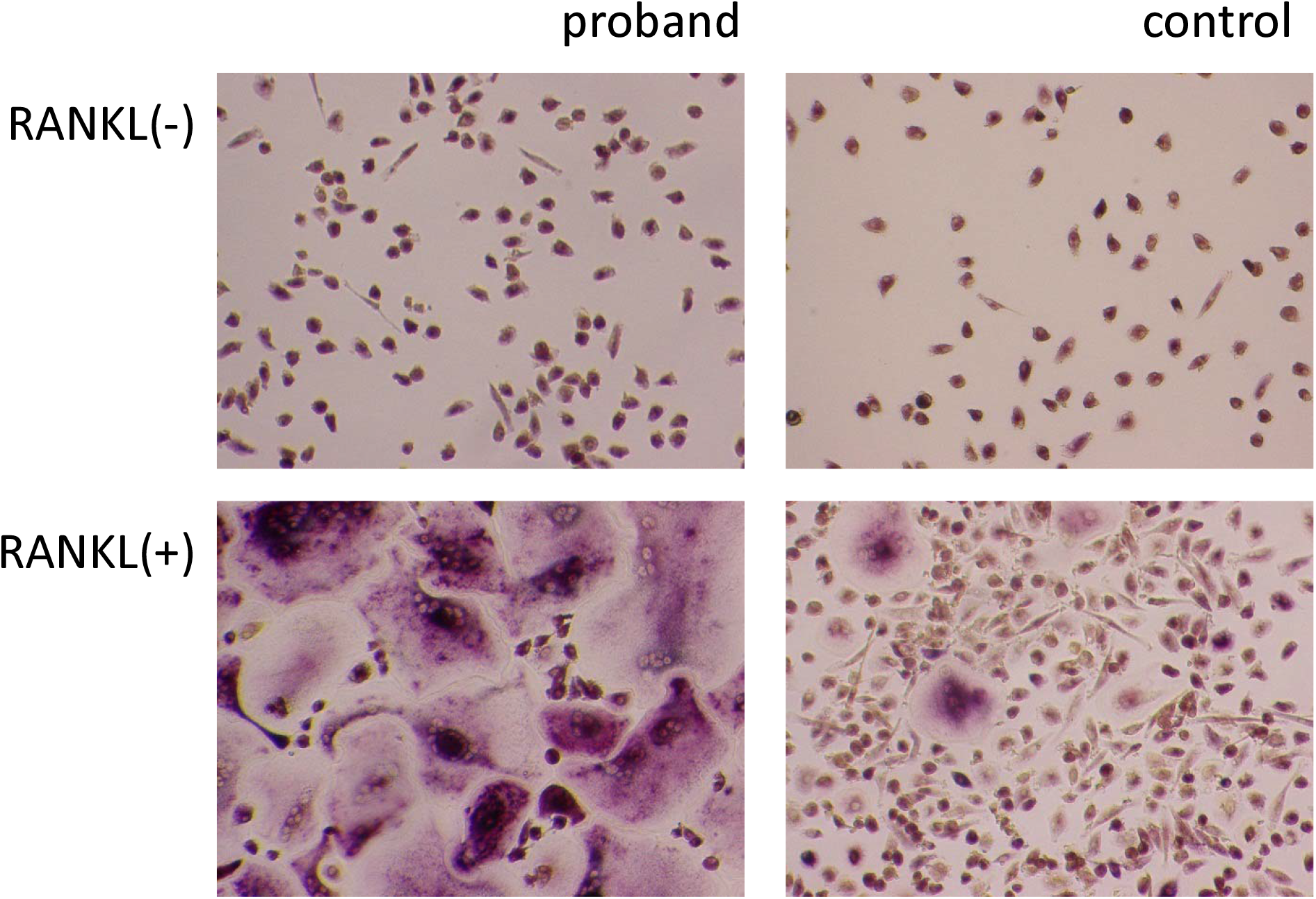
Circulating monocytes from the proband can differentiate into large TRAP-positive multinucleated osteoclast-like cells. Cells incubated for 4 days in the presence or absence of rhRANKL are shown, stained histochemically for TRAP.

## Notes

### Competing Interest Statement

The authors have declared no competing interest.

### Funding Statement

This work was supported by grants from The National Health and Medical Research Council of Australia (NHMRC: ID1047796), the AUVA (Austrian Social Insurance for Occupational Risk) and the OEGK (Austrian Social Health Insurance Fund). None of the authors or their institutions at any time received payment or services from any other third party for any aspect of the submitted work.

### Author Declarations

Royal Adelaide Hospital Human Research Ethics Committee (Approval No. RAH130114)

