## Supplementary Figure 1 for "A case of autosomal dominant osteopetrosis type II with a severe bone phenotype but no amino acid converting mutation in the *CLCN7* gene"

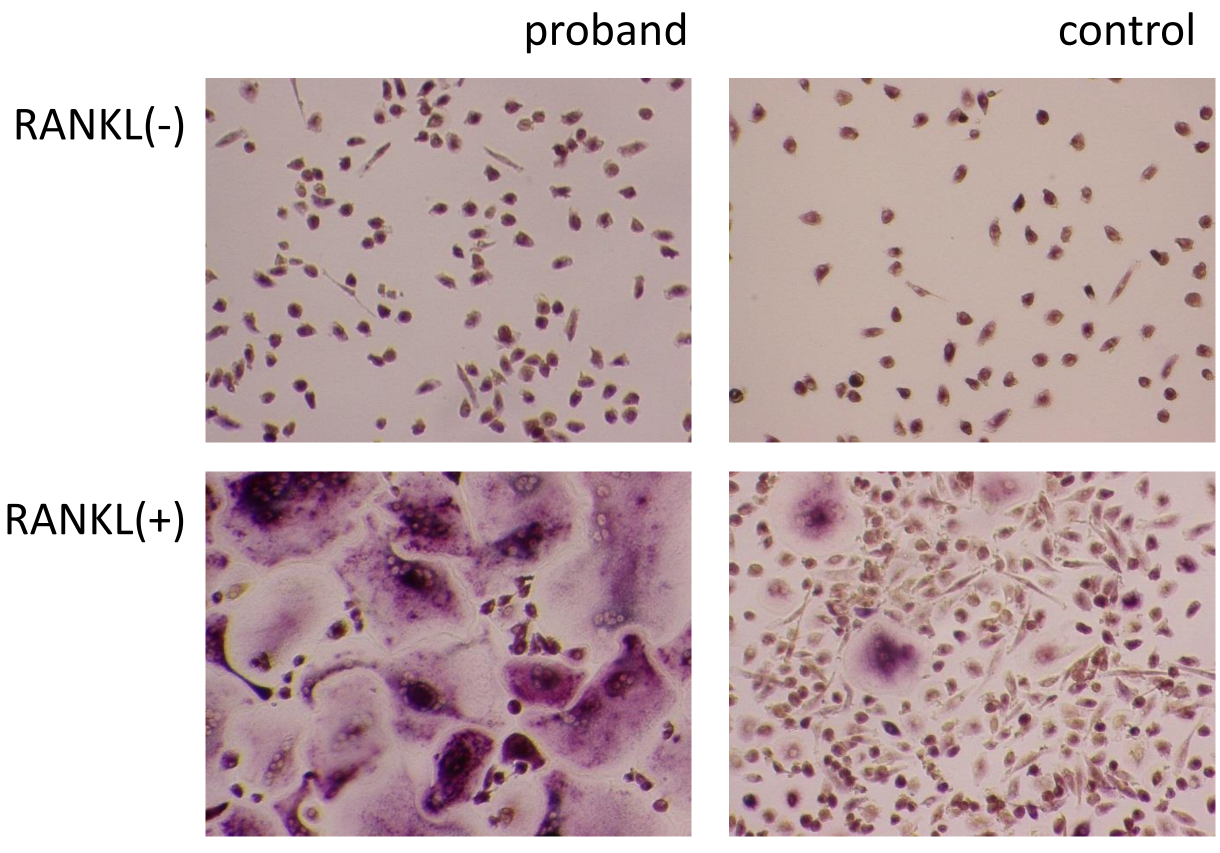


**Supplementary Fig. S1:** Circulating monocytes from the proband can differentiate into large TRAP-positive multinucleated osteoclast-like cells. Cells incubated for 4 days in the presence or absence of rhRANKL are shown, stained histochemically for TRAP.
